# Effective design of barrier enclosure to contain aerosol emissions from COVID-19 patients

**DOI:** 10.1101/2020.12.22.20246868

**Authors:** Dan Daniel, Marcus Lin, Irvan Luhung, Tony Lui, Anton Sadovoy, Xueqi Koh, Anqi Sng, Tuan Tran, Stephan C. Schuster, Xian Jun Loh, Oo Schwe Thet, Chee Keat Tan

## Abstract

Facing shortages of personal protective equipment, some clinicians have advocated the use of barrier enclosures (typically mounted over the head, with and without suction) to contain aerosol emissions from coronavirus disease 2019 (COVID-19) patients. There is however little evidence for its usefulness. To test the effectiveness of such a device, we built a manikin that can expire micron-sized aerosols at flow rates close to physiological conditions. We then placed the manikin inside the enclosure and used a laser sheet to visualize the aerosol leaking out. We show that with sufficient suction, it is possible to effectively contain aerosol from the manikin even at high flow rates (up to 60 L min^−1^) of oxygen, reducing aerosol exposure outside the enclosure by 99%. In contrast, a passive barrier without suction only reduces aerosol exposure by 60%.

---

The prolonged nature of the coronavirus disease 2019 (COVID-19) pandemic has resulted in global shortages of personal protective equipment (PPE), especially N95 respirators. As a result, some clinicians have resorted to using barrier enclosures typically mounted on the hospital bed over the patient’s head to contain any aerosol emissions, especially during aerosol generating procedures such as intubation [1–3]. Initial barrier design consisted of a passive enclosure without suction which can effectively stop large respiratory droplets [1], but not smaller aerosol droplets. It was later realized that active suction is critical to effectively contain micron-sized aerosol droplets [4–6]. However, the amount of suction required for effective aerosol containment remains unexplored, especially when the patient is subjected to treatment modalities involving high gas flow rates, e.g. high flow nasal cannula (HFNC) oxygen therapy. There is also little quantitative data for the reduction in aerosol exposure for barrier designs with and without suction.

Here, we used laser sheets to visualize the aerosol flow from a custom-built manikin expiring micron-sized water-glycerin droplets at flow rates close to physiological conditions [7]. We found that with sufficient suction, it is possible to effectively contain aerosol inside the enclosure even when the manikin is subjected to high flow rates (up to 60 L min^−1^) of supplementary oxygen. By spiking the water-glycerin droplets with fluorescein [8] and using spectrofluorometer to quantify the amount of fluorescein collected by air samplers outside the barrier, we were able to establish that a barrier enclosure with active suction can reduce potential aerosol exposure by more than 99%. In contrast, a passive barrier without suction only reduces aerosol exposure by 60%.

Given the growing evidence that COVID-19 is airborne and can spread through aerosol [9–11], a well-designed barrier enclosure can be useful in protecting healthcare workers from infectious COVID-19 patients.

## RESULTS AND DISCUSSIONS

The barrier enclosure was designed by local institutions of higher learning and used by a hospital in Singapore (See Supporting Figure S1 for detailed design). The physical enclosure (width, breadth and height of 92 cm × 60 cm × 70 cm) is made of perspex (Fig. 1a), with a plastic drape at the front, four openings (two at the headend and one on either side) for access to patients, and two suction ports (one on either side). Suction is provided by two wall units typically found in a hospital, each with a maximum suction rate of 60 L min^−1^. COVID-19 patients typically require oxygen therapy, with high flow nasal cannula (HFNC) oxygen therapy (Fig. 1b) showing positive medical outcomes [12–14]. The high oxygen flow rate (up to 60 L min^−1^) can however quickly disperse any bioaerosol over large distances, increasing the infection risk to healthcare workers. In this paper, we will show that aerosol can be effectively contained in the enclosure when the flow rate of the suction *Q*_suction_ exceeds the sum of the oxygen flow rate *Q*_O2_ and the expiration rate of the patient *Q*_air_ (Fig. 1c), i.e.

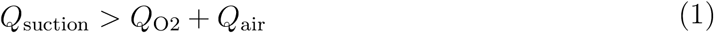

**FIG. 1.**
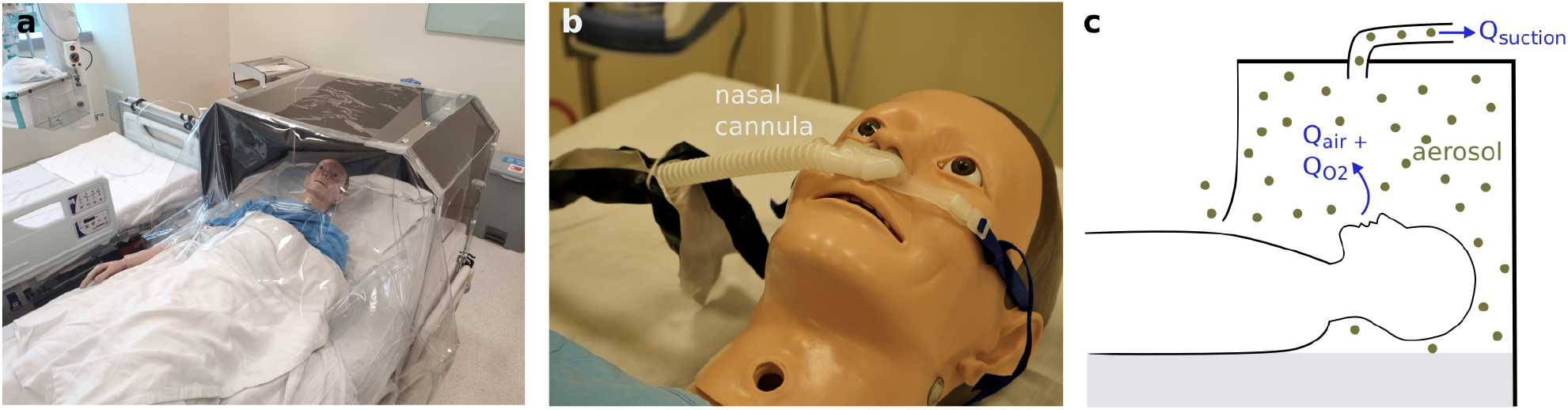
(a) Barrier enclosure used to contain aerosols from patients who typically require supplementary oxygen treatment (b) delivered through a nasal cannula. (c) For effective containment of aerosol (gray dots), the suction rate *Q*_suction_ must be higher than the oxygen flow rate *Q*_O2_ and the breathing (expiration) rate of the patient *Q*_air_ combined.

We first placed a custom-built manikin inside the barrier enclosure. Micron-sized water-glyerin aerosol droplets were generated inside a box (50 cm × 50 cm × 60 cm) using a fog machine (typically used in entertainment venues). See Supporting Fig S2 for droplet size distribution. Glycerin (50 v%) was added to water to prevent the aerosol droplets from evaporating. A manual resuscitator or an Ambu bag was then used to expel the aerosol from the box through the manikin into the barrier (Fig. 2a) once every 6 seconds for 20 minutes at flow rates close to human breath (0.63 L in 3 seconds or *Q*_air_ = 13 L min^−1^). The aerosol flow can be visualized by shining a blue laser sheet at the sagittal *z*–*y* plane (Fig. 2b and Supporting Video 1). At the same time, we shone a green laser sheet at the transverse *x*–*z* plane in front of the barrier to better visualize any leakage from the barrier enclosure.

**FIG. 2.**
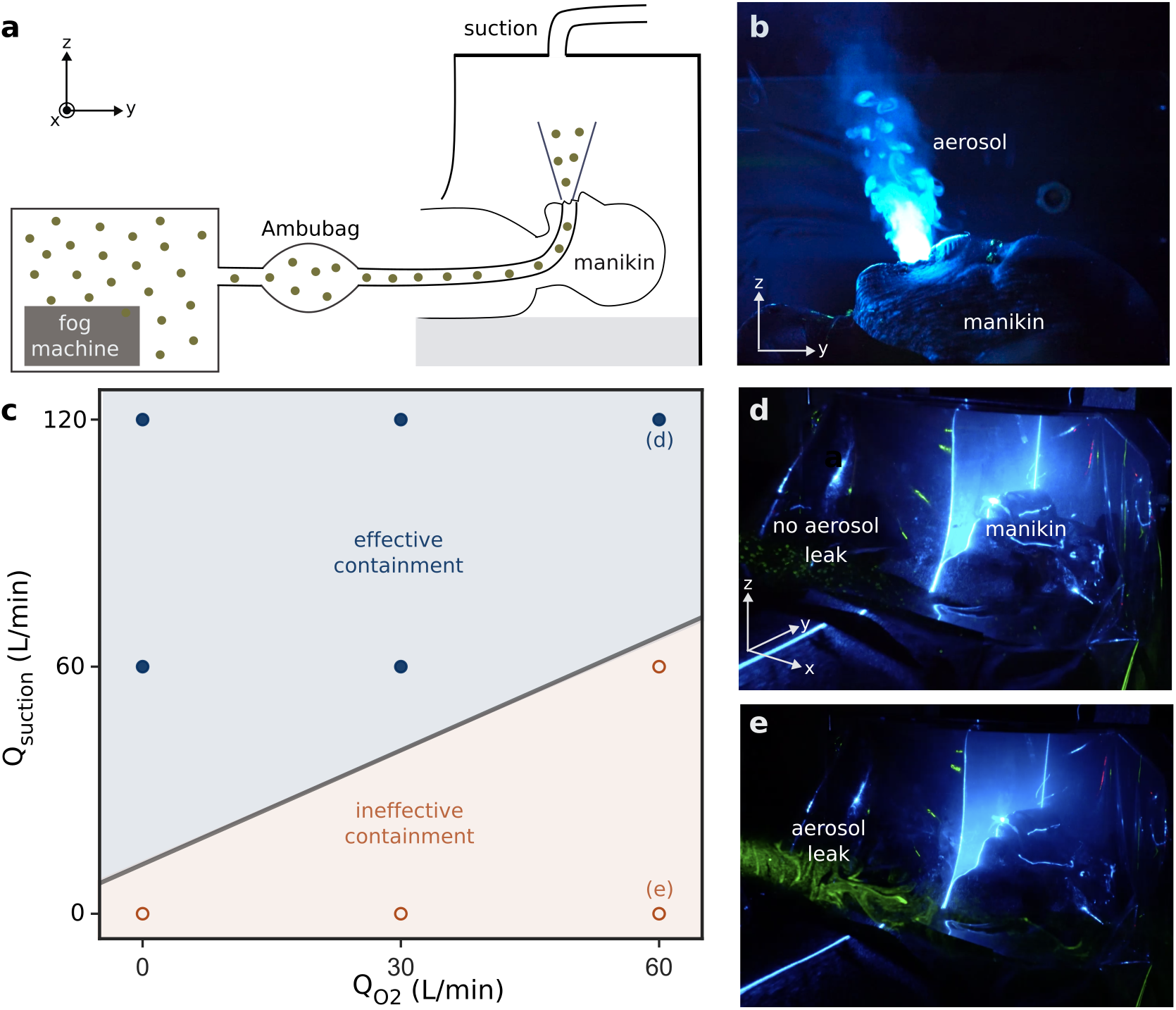
(a) Experimental setup used to simulate the breathing cycle. Nasal cannula used to deliver oxygen not shown. (b) Micron-sized glycerin-water aerosol droplets expelled from the manikin can be visualized by shining a blue laser sheet at the sagittal plane. (c) Effectiveness of aerosol containment depends on the applied suction rate *Q*_suction_ and the oxygen flow rate *Q*_O2_. Blue filled dots and red unfilled dots correspond to experimental conditions resulting in effective and ineffective containment, respectively. (d) An additional green laser sheet (transverse *x*–*z* plane) placed outside the enclosure is not visible in the absence of aerosol leak. (e) Aerosol leaking out from the enclosure scatters light and renders the green laser sheet visible. Experimental conditions corresponding to (d) and (e) are indicated in the phase diagram in (c).

To test the effectiveness of the aerosol containment under different conditions, we varied the suction rate *Q*_suction_ from 0 (no suction) to 60 and 120 L min^−1^ (1 and 2 wall suction units, respectively), while at the same time subjecting the manikin to a HFNC oxygen therapy at flow rates *Q*_O2_ = 0–60 L min^−1^ (Fig. 2c). Experimentally, we found that Equation 1 correctly predicts the criterion for effective aerosol confinement (The transition from effective to ineffective containment as predicted by equation 1 is indicated by the gray line in Figure 2c). For example, at maximal *Q*_O2_ = 60 L min^−1^ (and *Q*_air_ = 13 L min^−1^), two wall units with a combined *Q*_suction_ = 120 L min^−1^ are able to effectively confine the aerosol inside the barrier even after 20 minutes (Fig. 2d and Supporting Video 2). The green laser sheet is not visible since there is no aerosol to scatter the light and fresh air is continually being drawn in from gaps at the bottom of the plastic drape (dark area in Supporting Video 2 and Supporting Figure S3 is a region relatively free from aerosol). In contrast, with just one or no suction, the aerosol cannot be contained and spread quickly throughout the entire room (with a floor area of about 30 m^2^) within minutes. The aerosol leaking out scatters light strongly and renders the green laser sheet visible (Fig. 2e and Supporting Video 3) [15].

To assess the level of protection afforded by the barrier enclosure with and without suction, we added a small amount of fluorescein (0.5 g L^−1^) to the water-glycerin solution. The level of aerosol exposure can then be quantified by measuring the amount of fluorescein collected by filters of two air samplers (with a flow rate of 60 L min^−1^) placed 50 and 110 cm away from the barrier (samplers 1 and 2 in Figure 3a, respectively). The fluorescein trapped by the filter (Fig. 3b) can be dissolved in water (2 mL) and the amount deduced by spectrofluorometry.

**FIG. 3.**
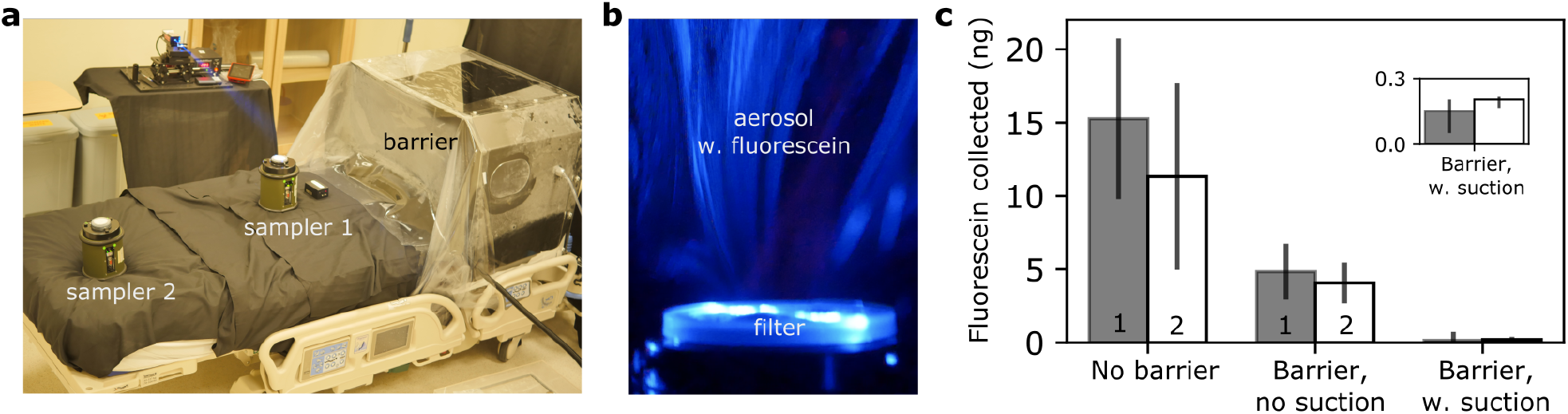
(a) To quantify the amount of aerosol leak, we placed two air samplers outside the barrier enclosure. (b) Aerosol droplets (with added fluorescein) were trapped by the filter on the air sampler, which can then be detected using spectrofluorometer. (c) The amount of fluorescein collected by air samplers 1 and 2 for enclosure barrier with and without suction can then be compared to the control, i.e. no barrier. Error bars are the standard deviation for triplicates.

We found that for HFNC oxygen therapy with no barrier at *Q*_O2_ = 60 L min^−1^, the amount of fluorescein collected after 20 minutes by samplers 1 and 2 are 15 ± 5 ng and 11 ± 6 ng, respectively (Fig. 3c). With a passive barrier and no suction (corresponding to case d in Figure 2c), the amount of fluorescein collected by samplers 1 and 2 were reduced by about 60% to 5 ± 2 and 4 ± 1 ng, respectively. At maximum suction of *Q*_suction_ = 120 L min^−1^ (corresponding to case e in Figure 2c), the amount of fluorescein reaching the two samplers were reduced by 99% to 0.15 ± 0.08 and 0.19 ± 0.01 ng.

## CONCLUSIONS

We have established the criterion for effective containment of aerosol for barrier enclosure, namely that the suction rate must exceed the oxygen flow rate and the expiration rate of the human breath. We show explicitly that for high flow (60 L min^−1^) nasal cannula oxygen therapy, it is possible to significantly reduce aerosol exposure outside the enclosure by 99% with sufficient suction (120 L min^−1^). Finally, the concept of barrier enclosure with active suction is very general and can easily be extended to settings outside of healthcare facilities, e.g. barrier enclosures can be installed on individual seats on a plane or train for safer travel during a pandemic.

## MATERIALS AND METHODS

### Simulating human breath cycle using a custom-built manikin

A 400 W fog machine, typically used in entertainment venues, was used to heat up and generate micron-sized aerosol from a glycerin-water mixture (50 v%) inside a 50 cm × 50 cm × 60 cm box within 3 seconds. See Supporting Figure S2 for the droplet size distribution. Food-grade glycerin was purhased from a baking shop. A manual resuscitator or an Ambu bag was then used to transfer the aerosol from the box through the mouth of the manikin into the barrier. The Ambu bag was compressed by hand and the volume pushed out during each compression cycle (0.63 ± 0.3 L) was determined by releasing the expelled air into a jar filled with water and measuring the volume of water displaced. The length of the compression cycle (3 s) was chosen to closely mimic the physiological parameters of the human breath. We did not find significant difference in the expelled volumes between different human operators. In a typical experiment, the Ambu bag was compressed for 3 seconds and released for another 3 seconds to mimic continuous breathing out of aerosol for 20 minutes. A metronome was used to help the operator keep in time.

### Fluorescein quantification

Aerosol (glycerol-water droplets with fluorescein) in the air was collected using a SASS 3100 Air Sampler from Research International running at a flow rate of 60 L min^−1^ for 20 min. SASS 3100 air sampler has been used previously to collect bioaerosols from the air [16]. The filter paper was then placed in 2 mL of deionized (DI) water inside a tube and shaken with a vortex mixer for 1 min to dissolve the trapped fluorescein. Fluorescein has an excitation and emission wavelengths of 490 and 514 nm, respectively. The fluorescein concentration (and hence the amount of fluorescein) in the 2 mL solution was then determined using a spectrofluorometer (Duetta, HORIBA scientific) by comparing its fluorescence intensity at 514 nm with those from calibration standards of known concentrations. The minimum concentration that can be measured using spectrofluorometer is 0.02 *µ*g L^−1^ or 0.04 ng in 2 mL solution.

## Data Availability

Data will be made available upon reasonable request.

## Authors’ contributions

D.D., L.X.J., C.K.T. and O.S.T. conceived the research idea and supervised the research. O.S.T. and C.K.T designed and built the barrier enclosure. D.D., M.L., T.L., A.S., X.K., T.T. contributed to the laser visualization experiment. I.L., S.S. and A.S. performed the air sampling and subsequent fluorescein quantification. D.D. and C.K.T. wrote up the manuscript. All authors have read and approved the manuscript.

## Supporting information

### I. LIST OF VIDEOS

Video 1: Manikin breathing out aerosol

Video 2: Effective containment of aerosol in the barrier enclosure

Video 3: Ineffective containment of aerosol in the barrier enclosure

### II. BARRIER ENCLOSURE DESIGN

**FIG. S1.**
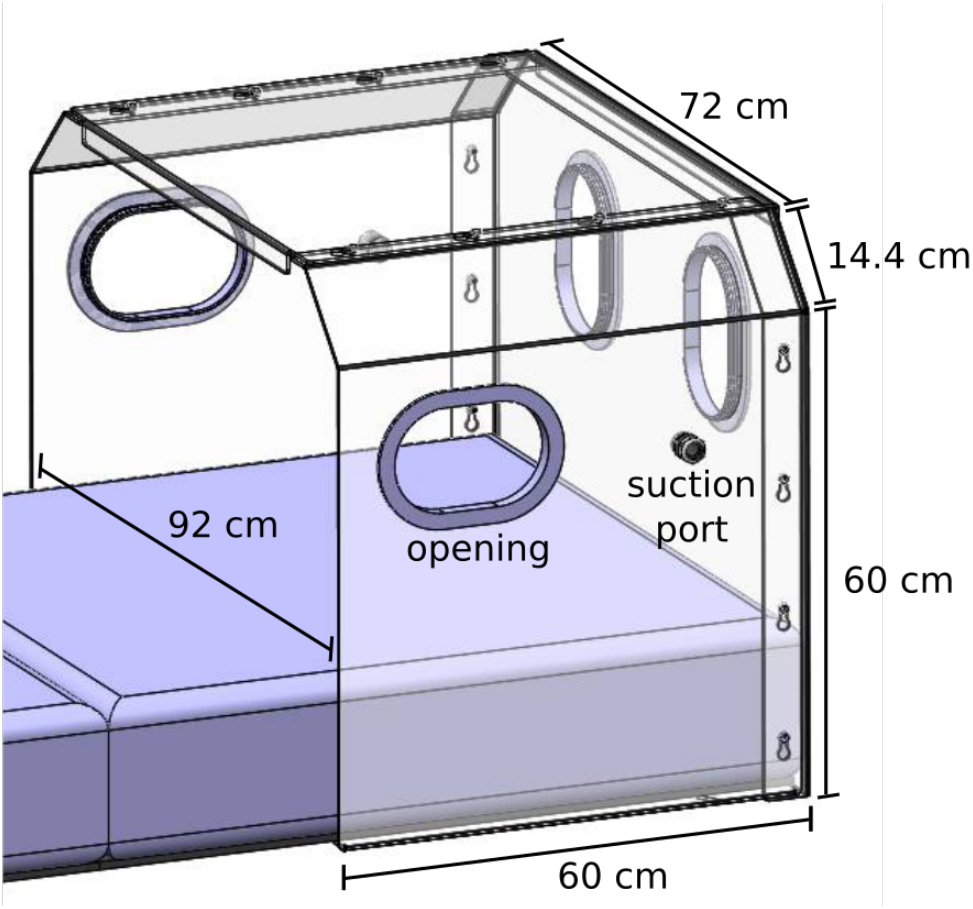
Barrier enclosure design

### III. DROPLET SIZE DISTRIBUTION

**FIG. S2.**
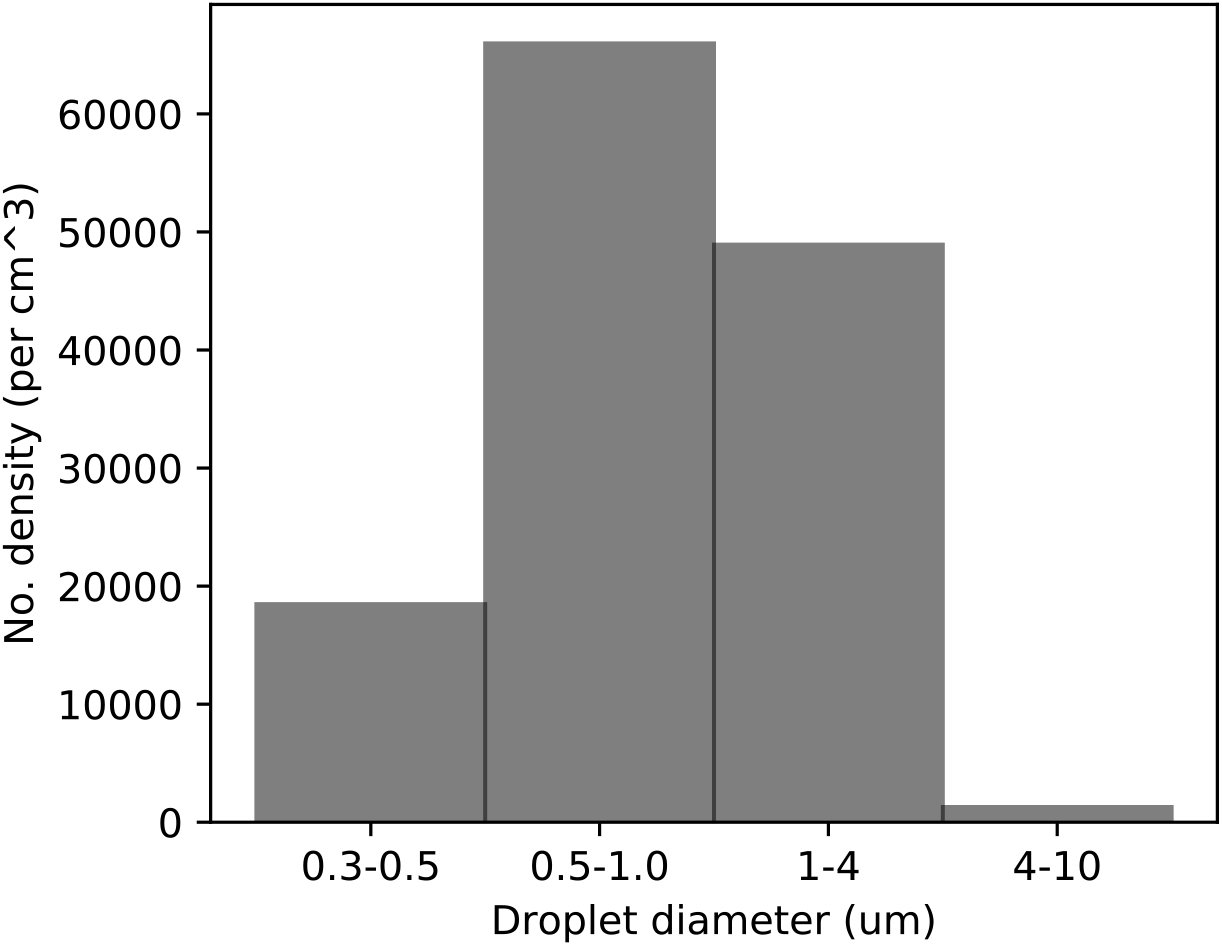
Droplet size distribution measured using an optical particle sizer.

### IV. VISUALIZATION OF AEROSOL FLOW

**FIG. S3.**
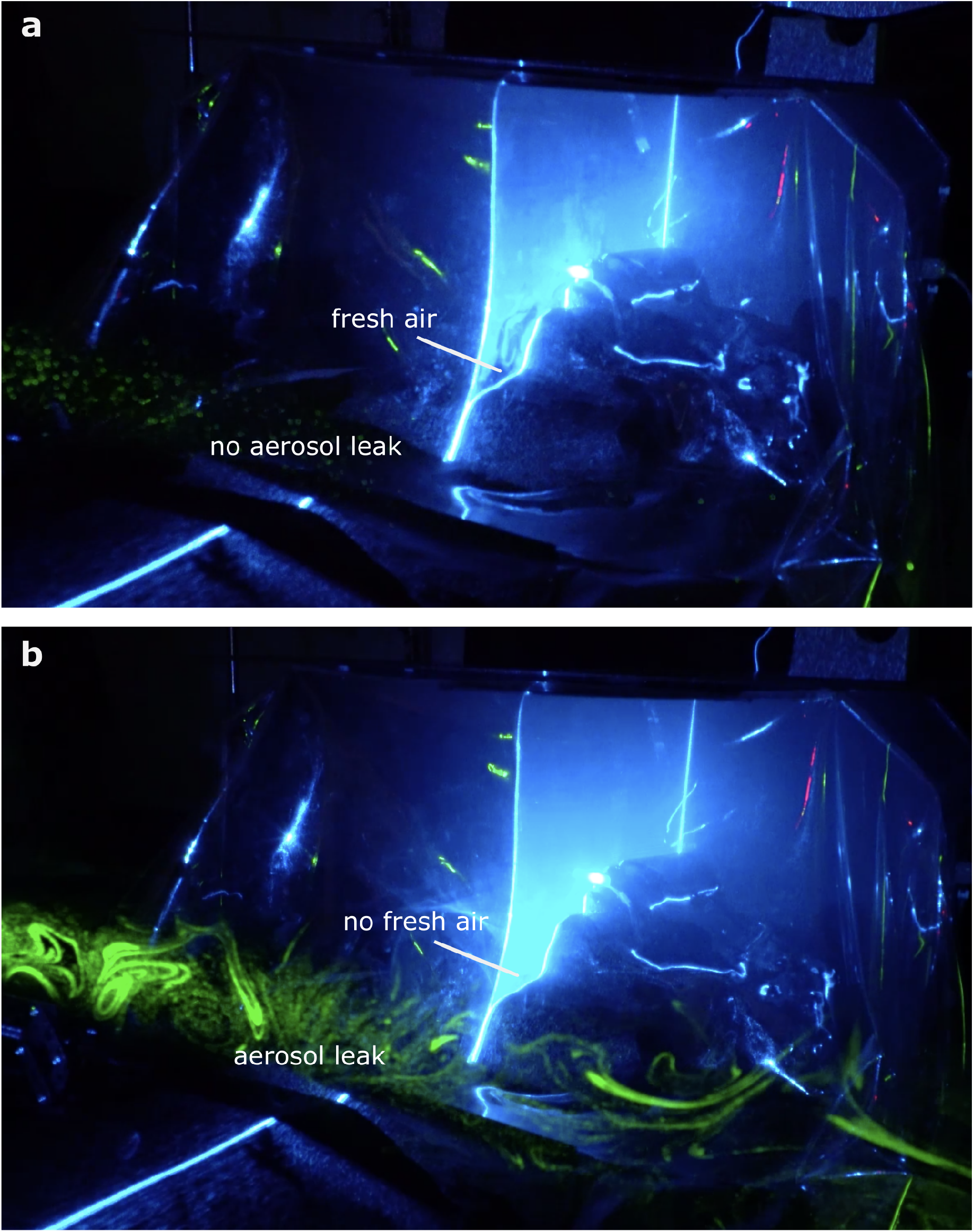
Visualization of aerosol flow (a) with and (b) without suction.

